# A Systematic Review of Polyherbal Plant Combinations in the Management of Type 2 Diabetes Mellitus in Africa

**DOI:** 10.1101/2025.10.16.25338054

**Authors:** Nokukhanya Thembane, Siphamandla Hlatshwayo, Sanele Mhlungu, Siboniso Sithole, Phikelelani Ngubane, Mlungisi Ngcobo, Nceba Gqaleni

## Abstract

This systematic review protocol outlines the planned approach to evaluate the efficacy and safety of polyherbal plant combinations in managing Type 2 Diabetes Mellitus (T2DM) within African populations. Given the rising burden of T2DM in Africa and limited access to conventional treatments, polyherbal formulations are widely used as alternative or complementary therapies. This review will include quantitative studies such as randomized controlled trials, cohort studies, and in vivo/in vitro research, as well as qualitative ethnopharmacological studies. Primary outcomes focus on glycemic control markers, fasting serum glucose, HbA1c, and postprandial glucose, while secondary outcomes include lipid profiles, safety, and phytochemical characteristics. Comprehensive literature searches will be conducted across multiple databases from 2011 to 2024. Risk of bias will be assessed, and where appropriate, meta-analyses will be performed. Findings aim to provide evidence-based insights to guide clinical practice, research, and policy on the use of polyherbal therapies for T2DM management in Africa.

## 4. Amendments

Any deviations or amendments to this protocol during the review process will be documented with the date, description, and rationale. Updates will be communicated by updating the PROSPERO record and noted in the final systematic review manuscript to maintain transparency and allow readers to track changes.

## 5. Support

Funding Source: None, there is no funder with any role in study design, data collection, analysis, interpretation, or publication.

## 6. Review Questions

- What is the efficacy of polyherbal plant combinations in managing Type 2 Diabetes Mellitus (T2DM) in African populations compared to single-plant preparations, placebo, or conventional antidiabetic drugs?
- What are the safety profiles and adverse effects of these polyherbal combinations?
- Which polyherbal combinations are most commonly used for T2DM in Africa?
- How do study designs vary among existing research on this topic?

## 7. Condition or Domain Being Studied

Type 2 Diabetes Mellitus (T2DM), with a focus on polyherbal medicinal plant combination interventions used within African populations.

## 8. Participants/Population

Inclusion Criteria:

- Adults (≥18 years) diagnosed with T2DM.
- Animal models of T2DM used in in vivo studies.
- Studies conducted in African countries or involving African populations.

Exclusion Criteria:

- Studies focusing solely on Type 1 Diabetes Mellitus.
- Monotherapy studies unless compared to polyherbal formulations.
- Studies not involving African populations or plants.

## 9. Interventions

Polyherbal formulations comprising two or more plant species used for the management of T2DM.

## 10. Comparators

Placebo, conventional antidiabetic drugs (e.g., metformin, insulin), single-plant monotherapies.

## 11. Types of Studies to Be Included

Quantitative Studies:

- Randomized Controlled Trials (RCTs)
- Non-randomized trials
- Cohort studies
- In vivo (animal) studies
- In vitro studies (for mechanistic insights)

Qualitative/Ethnopharmacological Studies:

- To provide contextual understanding of traditional use and formulation practices.

## 12. Outcomes

Primary Outcomes:

- Fasting Blood Glucose (FBG)
- Glycosylated Hemoglobin (HbA1c)
- Postprandial Glucose (PPG)

Secondary Outcomes:

- Lipid profile (Total cholesterol, LDL, HDL, triglycerides)
- Renal and liver function markers (e.g., serum creatinine, ALT, AST)
- Adverse events and safety profiles
- Phytochemical composition and standardization of polyherbal formulations

## 13. Search Strategy

Databases: MEDLINE (PubMed), Web of Science, Scopus, Google Scholar. Search Terms: “polyherbal,” “plant combinations,” “herbal medicine,” “traditional medicine,” “diabetes mellitus,” “Type 2 Diabetes,” “Africa,” and names of individual African countries. Language Restrictions: English. This review will include only studies published in English due to resource constraints for translation and to ensure accurate interpretation of study findings. While this may introduce some language bias, English is the predominant language of scientific publication in the region and the databases searched. Any potentially relevant non-English studies identified during screening will be documented and discussed in the limitations section. Time Frame: The date restrictions with 2011 - 2024; all available literature up to the date of search.

## 14. Data Extraction and Management

Two independent reviewers will screen titles, abstracts, and full texts. Data extraction will include:

- Study characteristics (author, year, country, design)
- Participant demographics
- Intervention and comparator details
- Outcome measures and results
- Adverse events
- Risk of bias assessments

Discrepancies will be resolved through discussion or by a third reviewer.

## 15. Risk of Bias Assessment

Risk of bias will be assessed using appropriate, established criteria tailored to the type of study. Where applicable, simplified or adapted tools will be used to evaluate methodological quality.

For example:

- Clinical studies will be assessed using a simplified version of the Cochrane Risk of Bias approach.
- Animal and in vitro studies will be evaluated based on key elements of study design such as randomization, blinding, and reporting transparency.
- The assessment will focus on identifying potential sources of bias and study limitations that may impact the reliability of the findings.

## 16. Data Synthesis

Quantitative Synthesis: Meta-analysis will be conducted if data are sufficiently homogenous using random-effects models.

Narrative Synthesis: For heterogeneous or qualitative data.

Subgroup Analyses: Based on type of comparator, study design, region/country, type of polyherbal formulation.

Heterogeneity Assessment: I^2^ statistic and Chi-square test. Publication Bias: Funnel plots and Egger’s test (if ≥10 studies).

## 17. Confidence in Cumulative Evidence

The overall strength of the body of evidence will be evaluated using a simplified approach based on GRADE principles. This will consider factors such as study design, consistency of results across studies, directness of evidence, and risk of bias. The assessment will provide a general indication of confidence in the findings, particularly for key outcomes related to efficacy and safety.

## 18. Ethics and Dissemination

Ethical Approval: Not required as this is a secondary analysis of published data. Dissemination Plan: Findings will be submitted to peer-reviewed journals and presented at relevant academic conferences.

## 5 Introduction

### 5.1 Rationale

Type 2 Diabetes Mellitus (T2DM) is a rapidly escalating public health challenge in sub-Saharan Africa, with profound impacts on individuals, communities, and strained healthcare systems. Recent evidence from the AWI-Gen longitudinal cohort study in four African countries (South Africa, Kenya, Ghana, Burkina Faso) shows that the prevalence of T2DM nearly doubled over six years, reaching 10.9% in adults aged 40 to 60 years (Chikwati et al., 2025). This alarming increase exceeds previous projections for the region and highlights the urgent need for effective interventions tailored to African populations. The International Diabetes Federation (IDF) estimates that currently 25 million adults in Africa live with diabetes, predominantly T2DM, and projects a staggering 142% increase by 2050 (IDF, 2024). Notably, more than half of people with diabetes in the WHO African Region remain undiagnosed or untreated, increasing their risk of severe complications such as cardiovascular and renal disease.

While conventional pharmacological treatments for T2DM exist, their accessibility and affordability in low-resource settings are limited. Additionally, long-term use is often associated with adverse side effects, necessitating alternative or complementary therapeutic options. Traditional medicine, particularly polyherbal plant combinations, is extensively used in Africa and may offer culturally accepted, cost-effective approaches to managing T2DM.Despite promising findings from studies on individual medicinal plants, there remains a lack of comprehensive synthesis regarding combined herbal formulations specifically for T2DM management in African contexts. This gap includes limited data on efficacy, safety, standardized dosages, and clinical outcomes.

This systematic review aims to fill this critical gap by evaluating existing evidence on the use of polyherbal combinations for T2DM management in Africa, focusing on glycaemic control, safety profiles, and usage patterns. The findings will inform future research priorities, policy development, and potential integration of effective traditional therapies into formal diabetes care frameworks.

### 5.2 Objectives

To systematically review and synthesize evidence on the efficacy and safety of polyherbal plant combinations for managing T2DM in African populations. Specifically:

- To assess glycemic control outcomes (FBG, HbA1c, PPG) associated with polyherbal therapies.
- To document safety profiles and adverse effects reported.
- To identify commonly used polyherbal combinations and study characteristics.

## 6. Methods

### 6.1 Eligibility Criteria

- Population: Adults (≥18 years) diagnosed with T2DM, and animal models relevant to T2DM from African contexts.
- Interventions: Polyherbal formulations containing two or more plant species aimed at managing T2DM.
- Comparators: Placebo, conventional antidiabetic drugs, or single-plant treatments.
- Outcomes: Primary glycemic control markers (FBG, HbA1c, PPG). Secondary—lipid profiles, safety parameters, renal/liver function, phytochemical constituents.
- Study Designs: Clinical trials (randomized and non-randomized), cohort studies, in vivo/in vitro studies, ethnopharmacological reports.
- Other criteria: English language restrictions; publication years [2011–2024].

### 6.2 Information Sources

- Databases to be searched include:
  - MEDLINE (PubMed)
  - Web of Science
  - Scopus
  - Google Scholar

Grey literature and trial registries will also be reviewed. The search will cover publications from 2011 up to 2024.

### 6.3 Search Strategy

A draft search strategy for PubMed includes: (polyherbal OR “plant combination” OR “herbal medicine” OR “traditional medicine”) AND (“type 2 diabetes” OR “T2DM”) AND (Africa) Filters for date and language will be applied as needed. The full search strategy will be adapted for each database. A draft search strategy for PubMed includes: (polyherbal OR “plant combination” OR “herbal medicine” OR “traditional medicine”) AND (“type 2 diabetes” OR “T2DM”) AND (Africa) Filters for date and language will be applied as needed. The full search strategy will be adapted for each database.

### 6.4 Study Records

#### Data Management

All records will be managed using reference software (e.g., EndNote or Zotero). Duplicates will be removed prior to screening.

#### Selection Process

Two independent reviewers will screen titles and abstracts for eligibility. Full texts of potentially relevant articles will be assessed against inclusion criteria. Discrepancies will be resolved by discussion or third reviewer arbitration.

#### Data Collection Process

Data extraction will be performed independently by two reviewers using a piloted standardized form. Disagreements will be resolved by consensus.

### 6.5 Data Items

- Data to be extracted include:
  - Study characteristics (authors, year, country, design)
  - Population details (sample size, demographics)
  - Intervention specifics (plants used, dosages, duration)
  - Comparator details
  - Outcomes (glycemic markers, lipid profiles, safety/adverse effects)
  - Funding sources and conflicts of interest
  - Risk of bias indicators

### 6.6 Outcomes and Prioritization

- Primary outcomes: Glycemic control (FBG, HbA1c, PPG) due to direct relevance to diabetes management.
- Secondary outcomes: Lipid profiles, safety/adverse events, renal and liver function markers, phytochemical composition.

### 6.7 Risk of Bias in Individual Studies

Assessment will be conducted independently by two reviewers, with conflicts resolved by discussion.

### 6.8 Data Synthesis

If studies are sufficiently homogenous, a meta-analysis using random-effects models will be performed. Summary measures will include mean differences or standardized mean differences for continuous outcomes. Statistical heterogeneity will be assessed via I^2^ statistics. If meta-analysis is not feasible, a narrative synthesis will be conducted, organized around intervention types and outcomes.

### 6.9 Meta-bias(es)

Publication bias will be evaluated by funnel plots and Egger’s test if ≥10 studies are included in meta-analyses. Selective reporting will be assessed by comparing protocols (if available) to published results.

### 6.10 Confidence in Cumulative Evidence

The strength of evidence will be assessed using the GRADE approach, considering risk of bias, inconsistency, indirectness, imprecision, and publication bias.

## Data Availability

All data referred to in this systematic review protocol are from publicly available sources and will be fully reported in the final review.

## Author Contributions

| Task | Duration |
| --- | --- |
| Protocol Development | 24 May 2025 |
| Literature Search |  |
| Screening |  |
| Data Extraction |  |
| Risk of Bias Assessment |  |
| Data Synthesis |  |
| Manuscript Drafting |  |

### Guarantor

- **Nokukhanya Thembane (N.T.)** is the guarantor of the review and accepts full responsibility for the integrity of the work.

## Notes

Any deviations from this protocol will be documented and justified in the final review manuscript.

## Timeline Overview

The review will commence after protocol completion and before data extraction begins

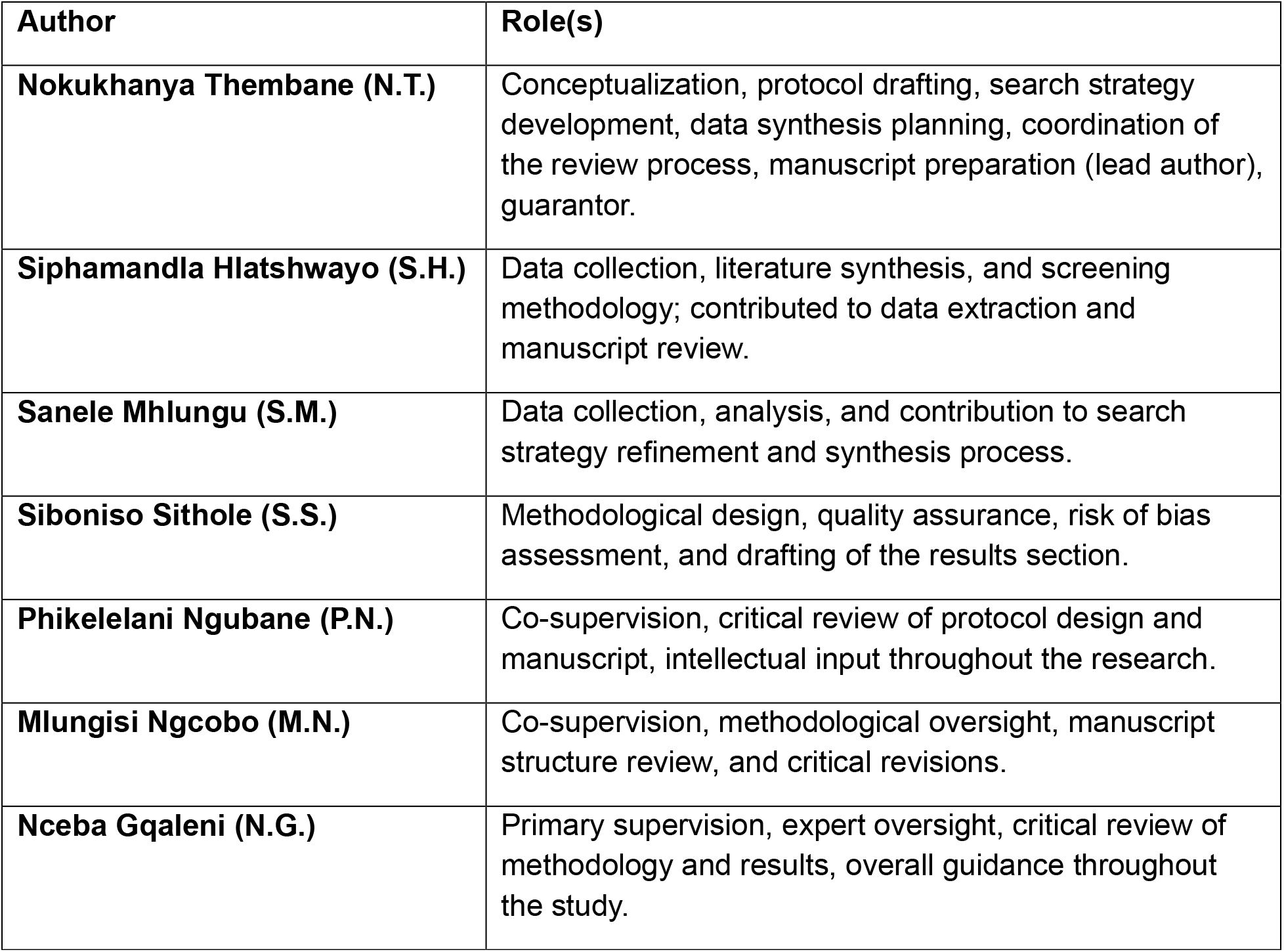

**End date:** 30 November 2025

**(This is the expected date for the review’s completion, including manuscript finalization.)**

## Notes

### Competing Interest Statement

The authors have declared no competing interest.

### Funding Statement

This study did not receive any funding.

### Author Declarations

The study used ONLY openly available human data originally located at: PubMed, Scopus, Google Scholar

